# Genetic liability for anxiety associates with treatment response to the monoamine stabilizer OSU6162 in alcohol dependence

**DOI:** 10.1101/2024.03.11.24304098

**Authors:** Mun-Gwan Hong, Lotfi Khemiri, Joar Guterstam, Johan Franck, Nitya Jayaram-Lindström, Philippe A. Melas

## Abstract

OSU6162, a monoamine stabilizer, has demonstrated efficacy in reducing alcohol and anxiety-related behaviors in preclinical settings. In a previous randomized, double-blind, placebo-controlled trial involving patients with alcohol dependence (AD), we found that OSU6162 significantly reduced craving for alcohol, but did not alter drinking behaviors. In the present study, we explored the hypothesis that genetic predispositions related to AD or associated traits, might influence the response to OSU6162 treatment in original trial participants (N=56). To investigate this, we calculated polygenic risk scores (PRSs) over several statistical significance thresholds from genome-wide association studies on (i) alcohol use disorder and alcohol consumption (N=200-202k), (ii) problematic alcohol use (N=435k), (iii) drinks per week (N=666k), (iv) major depression (N=500k), and (v) anxiety (using both case-control comparisons and quantitative anxiety factor scores, N=17-18k). Linear regression analyses assessing the interaction effects between PRSs and treatment type (OSU6162 or placebo) identified significant associations when considering anxiety factor scores (FDR<0.05). Specifically, in OSU6162-treated AD individuals, there was a negative correlation between anxiety factor PRS (at the genome-wide significance threshold that included one genetic variant) and several drinking outcomes, including number of drinks consumed, percentage of heavy drinking days, and changes in blood phosphatidylethanol (PEth) levels. These correlations were absent in the placebo group. While preliminary, these findings suggest the potential utility of anxiety PRS in predicting response to OSU6162 treatment in AD. Further research using larger cohorts and more comprehensive genetic data is necessary to confirm these results and to advance personalized medicine approaches in alcohol use disorder.

## Introduction

Alcohol dependence (AD) is a complex disorder characterized by dysregulated dopaminergic and serotonergic brain systems, which are crucial in modulating reward, craving, and cognitive functions (Sari et al., 2011; Soderpalm and Ericson, 2013). AD is also often associated with co-occurring anxiety and depressive disorders, further complicating the clinical picture (Burns and Teesson, 2002). These comorbid conditions share underlying familial liabilities with substance misuse (Virtanen et al., 2020) and have been linked to disturbances in monoamine neurotransmitter signaling (Belujon and Grace, 2017; DeGroot et al., 2020; Lawther et al., 2020). The compound (-)-OSU6162 (OSU6162), also known as PNU-96391, has emerged as a promising candidate in addressing these neurochemical imbalances.

OSU6162 stabilizes dopaminergic and serotonergic signaling pathways by acting as a neutral antagonist or a weak partial agonist at dopamine D2 and serotonin 5-HT2A receptors, (Burstein et al., 2011; Carlsson et al., 2011; Dyhring et al., 2010; Tolboom et al., 2015). Its documented efficacy in normalizing psychomotor activity and striatal dopaminergic function (Rung et al., 2008; Tedroff et al., 1998) formed the basis for its application in disorders marked by dopaminergic dysregulation, such as AD. Preclinical studies in long-term drinking rats have shown that OSU6162 reduces voluntary alcohol consumption, withdrawal symptoms, and the reinstatement of alcohol seeking (Fredriksson et al., 2019; Steensland et al., 2012), alongside mitigating anxiety-like behaviors (Melas et al., 2021) and correcting downregulated dopamine output in the nucleus accumbens (Feltmann et al., 2016).

In a Phase II human study, the efficacy of OSU6162 on drinking, craving, and mood in AD individuals was evaluated (Khemiri et al., 2015). The treatment was safe, well-tolerated, and notably reduced priming-induced craving and the subjective liking of alcohol. Additionally, OSU6162 improved certain cognitive functions, including future planning, verbal divergent thinking, and emotional recognition speed (Khemiri et al., 2019). Despite these promising findings, the study found no evidence of any treatment effects on alcohol intake (Khemiri et al., 2015). However, the study had a short duration, and there have not yet been any long-term clinical trials assessing the potential effects of OSU6162 on drinking outcomes.

In the present study, we aim to investigate whether genetic predispositions, particularly related to AD and the comorbid disorders of anxiety and depression, may influence therapeutic responses to OSU6162 treatment. By leveraging genome-wide association study (GWAS) data to calculate polygenic risk scores (PRSs) for alcohol use disorder, problematic alcohol use, alcohol consumption, major depressive disorder, and anxiety, the aim was to uncover putative genetic underpinnings of treatment response. This approach could benefit personalized medicine efforts in AD treatment, particularly when utilizing monoamine stabilizers like OSU6162, which could be tailored to individual genetic profiles.

## Methods

### Study design and participants

This study extends upon a previous randomized controlled trial with a double-blind, placebo-controlled design, where we enrolled 56 alcohol-dependent (AD) male and female participants, who were randomized to receive either OSU6162 (N=28) or placebo (N=28) for a 14-day period. For complete details on methods see (Khemiri et al., 2015). In brief, the study included three follow-up visits within the 14-day treatment period and a laboratory-based alcohol craving test session on day 15. Follow-up visits encompassed electrocardiogram (ECG), blood and urine sample collection, medication dispensing, breathalyzer tests, and self-reported drinking, mood, and adverse events. Participants, aged 20 to 55 years, met DSM-IV criteria for alcohol dependence, reported at least 45 heavy drinking days (HDD) in the preceding 90 days, and abstained from alcohol for 4 to 14 days before inclusion, confirmed by Timeline Follow Back (TLFB) interview (Sobell and Sobell, 1992) and breathalyzer. Exclusion criteria included other substance use disorders (except nicotine), schizophrenia, bipolar disorder, major depression, and significant cardiac or ECG abnormalities. The study was conducted in accordance with Good Clinical Practice and the Declaration of Helsinki, receiving approval from the regional ethical review board in Stockholm and the Swedish Medical Products Agency, and was registered in the European Clinical Trials Database (EudraCT; 2011-003133-34). All participants provided written informed consent.

### Clinical measures and alcohol craving test sessions

Mood and craving were assessed using the Montgomery-Åsberg Depression Self-Rating Scale (MADRS-S) (Svanborg and Asberg, 2001) and the Penn Alcohol Craving Scale (PACS) (Flannery et al., 1999), respectively. Alcohol consumption was quantified through changes in percent HDD, percent drinking days, and phosphatidylethanol (PEth) serum levels, along with the number of drinks, and percentage of both drinking days and HDD during the 14-day treatment period. On day 15, laboratory-based alcohol craving test sessions were conducted based on Hammarberg et al. (Hammarberg et al., 2009), involving three sessions triggered by alcohol-specific cues, neutral stimuli, and a priming dose of alcohol. Craving was evaluated using the shortened Swedish version of the Desire for Alcohol Questionnaire (Short-DAQ) (Love et al., 1998) and the Visual Analog Scale (VAS).

### GWAS data and quality control steps

AD patient genotypes were determined using the Illumina Infinium Global Screening Array-24 v2.0 (Illumina Inc., San Diego, CA, USA). Post-quality control (QC), 547,984 out of 730,059 genetic variants were retained for analysis, filtering out variants with over 10% missing genotypes, minor allele count of one or none, or significant Hardy-Weinberg equilibrium violation (P < 1e-10).

### Polygenic risk scores

PRSs were derived from seven base datasets, including GWASs for alcohol use disorder (Kranzler et al., 2019), alcohol consumption based on Alcohol Use Disorder Identification Test-Consumption (AUDIT-C) scores (Kranzler et al., 2019), problematic alcohol use (Zhou et al., 2020), drinks per week (Saunders et al., 2022), major depression (Howard et al., 2019), and two approaches to anxiety disorder phenotyping (case-control comparisons and quantitative anxiety factor scores) (Otowa et al., 2016). The details of the base datasets, including number of subjects and ancestries, are shown in Table S1. The PRSs were computed using PRSice-2 software v. 2.3.5 (Choi and O ‘Reilly, 2019) and the formula:

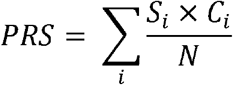

where *S*_*i*_ is the summary statistic for genetic variant *i* from the base dataset, *C*_*i*_ the observed effect-allele count of the variant *i* in the target dataset, and *N* the total number of alleles included in the PRS computation. The software was set to generate nine PRSs per risk for different p-value cutoffs from genome-wide significance to no-association, i.e., 5e-08, 0.001, 0.05, 0.1, 0.2, 0.3, 0.4, 0.5 and 1, excluding variants on sex chromosomes. Default software settings were maintained for other parameters, including a clumping threshold set at P ≤ 1, an r-squared value ≥ 0.1, and a maximum distance between SNPs of 250kb.

### Statistical analyses and bioinformatic environment

Data handling and analyses were conducted using R version 4.2.2 (2022-10-31) and the tidyverse (v. 2.0.0) package. To assess the associations between PRSs and clinical outcomes, we employed linear regression models. Specifically, we tested the interaction effects of treatment (OSU6162 or placebo) with PRS on the clinical measures. Multiple testing correction was applied using the Benjamini-Hochberg method to control the False Discovery Rate (FDR) at 5%. The strength and significance of the relationship between PRS and clinical response was also quantified using t-statistics for the estimated regression slopes.

## Results

### Genetic variants in PRS calculations for disorders or traits

As indicated in Table 1, the base GWAS datasets provided summary statistics for over 6 million genetic variants of European or mixed ancestry cohorts across seven disorders or traits, including alcohol use disorder, alcohol consumption (AUDIT-C), problematic alcohol use, drinks per week, depression, and anxiety disorder (binary and continuous). Matching reference SNP IDs between the base datasets and our study (target) cohort resulted in a common variant range of 256,268 – 499,638, representing approximately 47% to 91% of the QC-passed genetic variants. Due to linkage disequilibrium, the number of variants used for PRS calculations was reduced to 85,590 – 191,055 through clumping.

**Table 1.** Summary of statistical measures and variant counts per disorder or trait in each GWAS.

| Disorder or trait | Stat | Nr of variants |  |  |
| --- | --- | --- | --- | --- |
|  |  | in base dataset | in both base and target dataset<sup>‡</sup> | after clumping |
| Anxiety (case-control) | Odds ratio | 6,330,995 | 281,394 | 86,684 |
| Anxiety (factor score) | $\beta$ | 6,306,612 | 279,053 | 85,590 |
| Alcohol use disorder | Odds ratio | 6,895,250 | 256,268 | 110,604 |
| Alcohol consumption (AUDIT-C) | $\beta$ | 6,898,149 | 256,452 | 110,779 |
| Depression | Odds ratio | 8,483,301 | 448,812 | 171,188 |
| Drinks per week | $\beta$ | 13,267,983 | 499,638 | 191,055 |
| Problematic alcohol use | $\beta$ | 14,068,117 | 478,784 | 183,689 |

\*Number of variants shared between the base and target datasets, retained for subsequent clumping analysis. AUDIT-C: Alcohol Use Disorder Identification Test-Consumption

### Genetic liability for anxiety correlates with treatment response to OSU6162

Initial analyses revealed no significant differences in PRSs between the OSU6162 and placebo groups (Table S2). Subsequent linear models examined the interaction between treatment and PRSs across fourteen clinical measures with varying p-value cutoffs. No significant interactions were found for PRSs related to alcohol use disorder, alcohol consumption (AUDIT-C), problematic alcohol use, drinks per week, and anxiety characterized as binary outcomes (Tables S3-S7). However, significant interactions were observed between treatment and the anxiety factor score PRS at the genome-wide significance level. Notably, this interaction associated with the number of drinks consumed, percentage of HDD, percentage of drinking days, and changes in blood phosphatidylethanol (PEth) levels (FDR<0.05; in bold, Table 2). A trend was also observed for the change in the percentage of drinking days before and after the trial (FDR=0.06; in bold, Table 2). Figure 1 illustrates these findings, highlighting a robust negative correlation between anxiety PRS and drinking metrics in the OSU6162-treated group, as opposed to negligible correlations in the placebo group (t-statistics for the slopes of OSU6162-treatment vs. placebo: Diff_Peth: -3.06 vs. 0.17, P=0.0057 vs. P=0.86; TLFB_study_drinks: -3.85 vs. 1.89, P=0.00086 vs. P=0.071; TLFB_study_perc_HDD: -3.84 vs. 1.00, P=0.00090 vs. P=0.33; TLFB_study_perc_drinkingdays: -4.86 vs. 1.43, P=0.000074 vs. P=0.17). This suggests that individuals with higher genetic predisposition for anxiety may drink less when treated with OSU6162. It is, however, important to note that these observations are based on a single genome-wide significant SNP (rs1067327) for the anxiety factor score PRS. Finally, in examining PRS for depression with all genetic variants included (p-value cutoff=1), a weak significance emerged in one craving session (DAQ – active cue, FDR=0.04; in bold, Table S8), with higher PRSs inversely related to craving in the placebo group (Fig. S1; t-statistic for the slope of OSU6162-treatment vs. placebo: 1.26 vs. -3.22, P=0.22 vs. P=0.0041). However, this significance did not extend to the second active cue session (VAS – active cue, FDR=0.75, Table S8) nor to other cue sessions (e.g., priming), and it was not observed in the OSU6162 group (Table S8). This isolated significance suggests the potential for a type I error in the DAQ association.

**Table 2.**
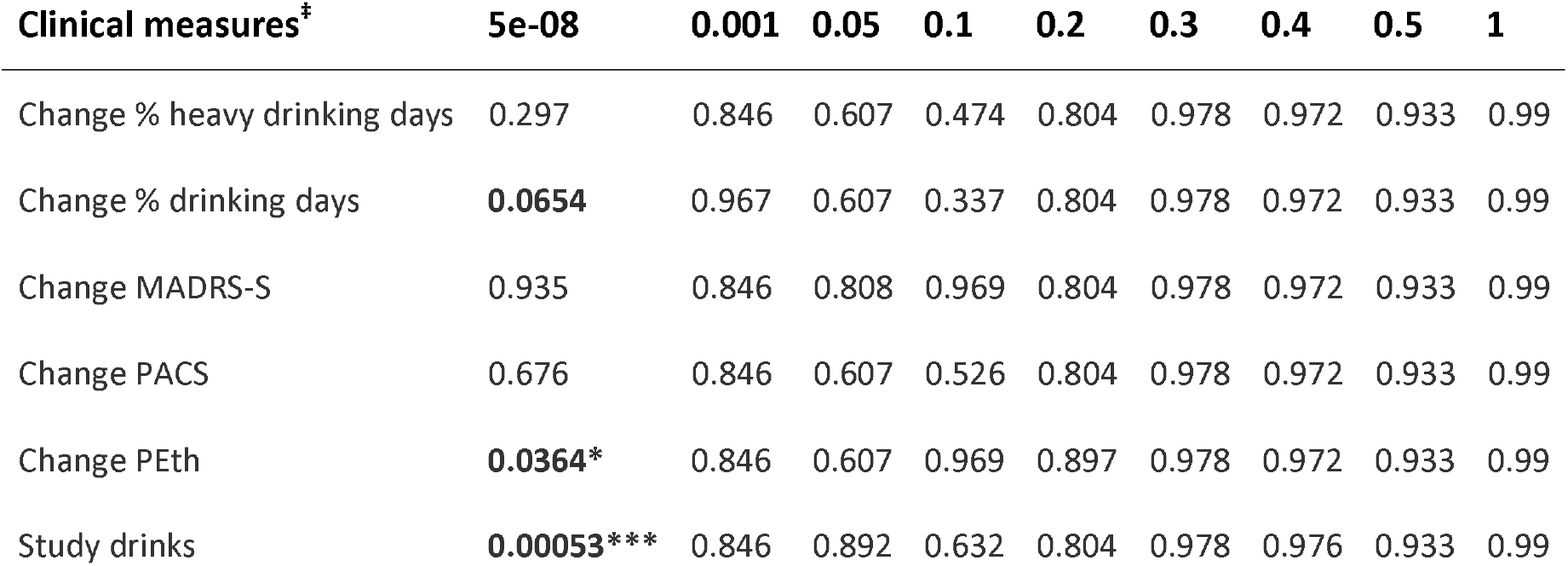
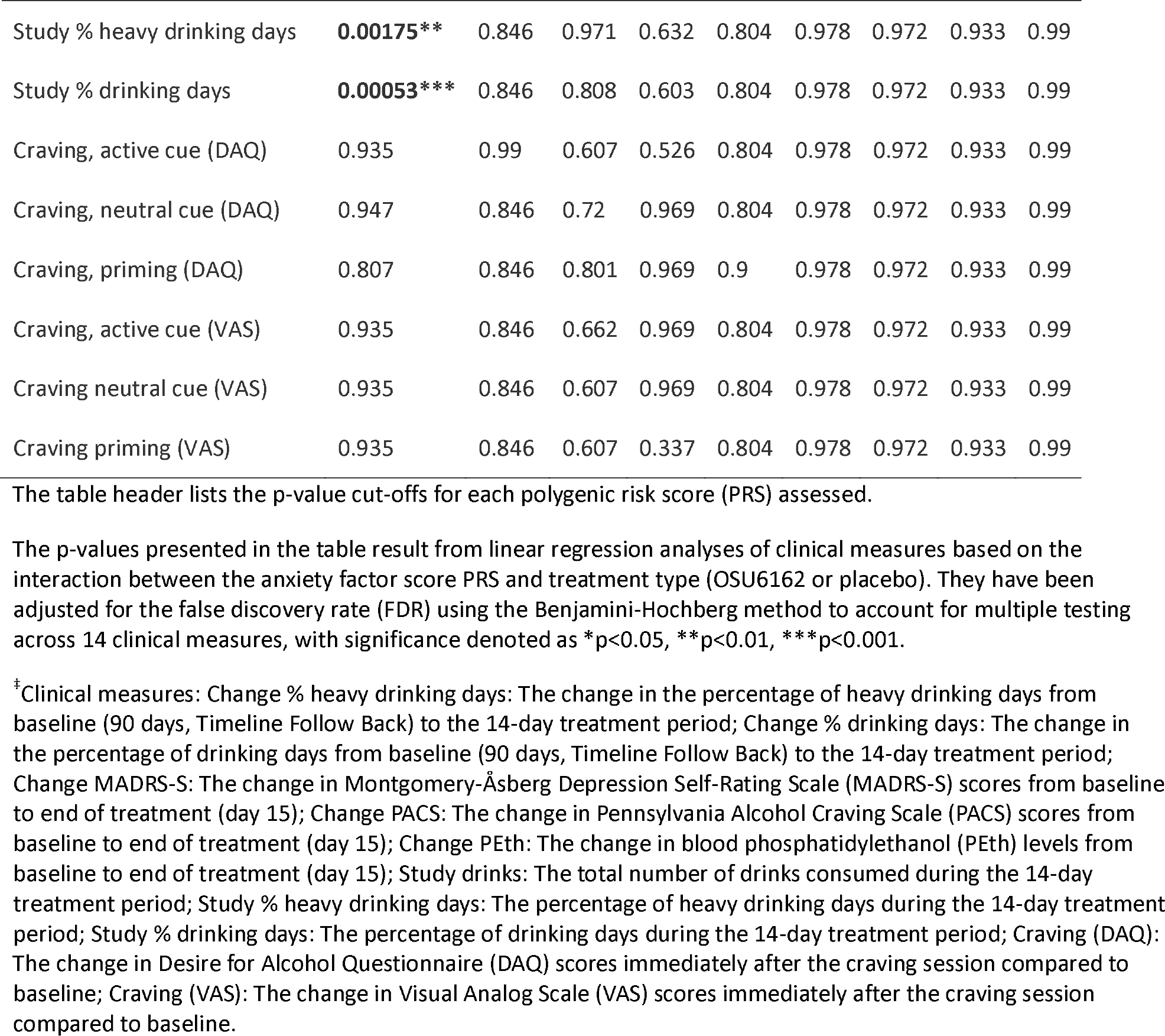
Impact of anxiety factor score PRS on clinical measures: Interaction with OSU6162 or placebo treatment.

**Fig 1.**
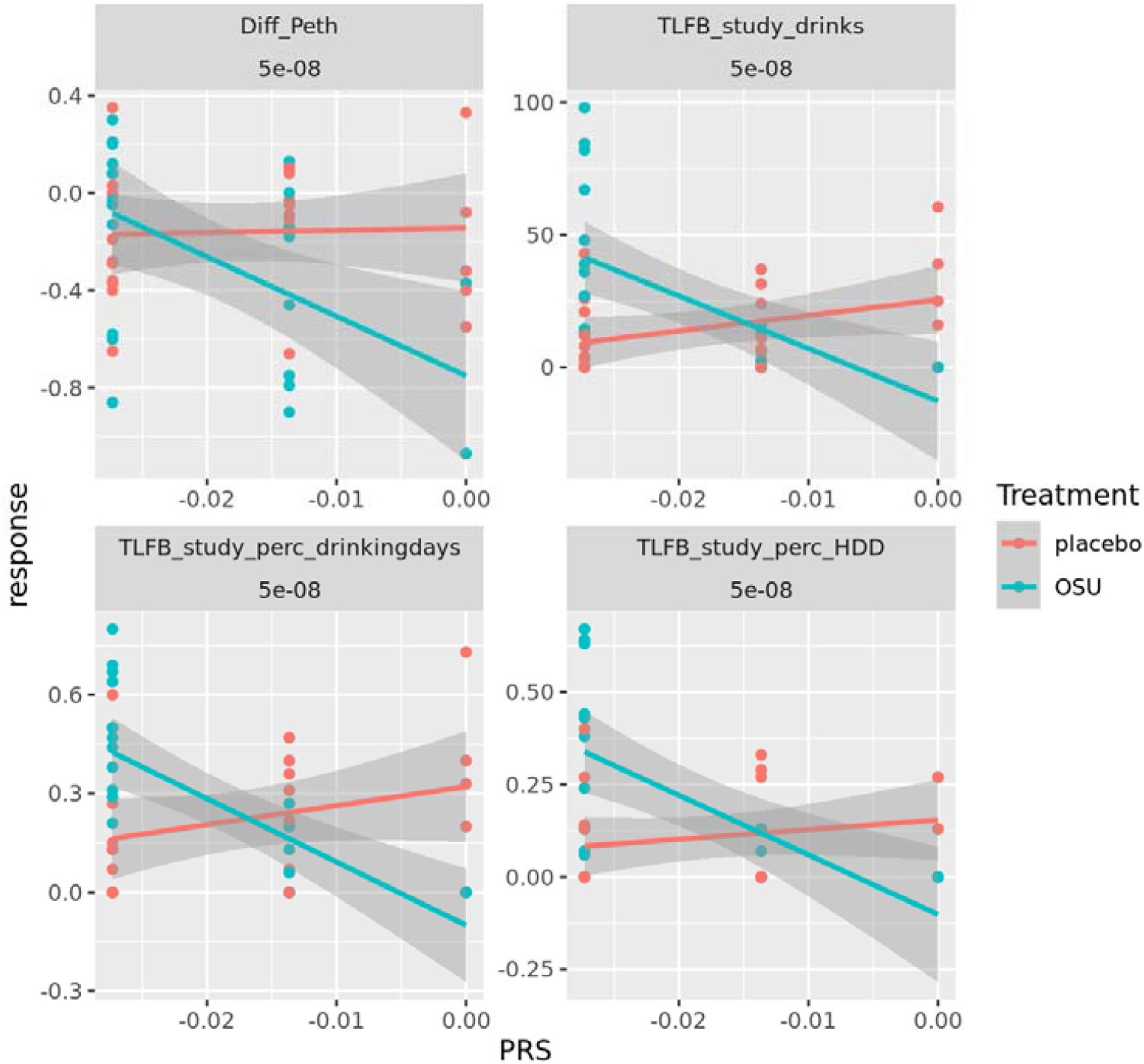
Correlations between anxiety factor PRS and clinical outcomes in alcohol dependent individuals on OSU6162 or placebo. In OSU6162-treated AD patients, a negative correlation was found between the anxiety factor PRS (at the genome-wide significance threshold that included one SNP) and measures of alcohol consumption during the trial, specifically: the number of drinks consumed, the percentage of drinking days, the percentage of heavy drinking days (HDD), and changes in blood phosphatidylethanol (PEth) levels post-treatment. These associations were not present in the placebo group. Diff_Peth: The change in blood phosphatidylethanol (PEth) levels from baseline to end of treatment (day 15); TLFB_study_drinks: The total number of drinks consumed during the 14-day treatment period; TLFB_study_perc_drinkingdays: The percentage of drinking days during the 14-day treatment period; TLFB_study_perc_HDD: The percentage of heavy drinking days during the 14-day treatment period; 5e-08: The p-value cutoff for PRS; TLFB: Timeline Follow Back. Treatment groups are indicated by color: red for placebo and teal for OSU6162 (OSU).

## Discussion

Despite AD presenting a significant global health burden, a substantial number of affected individuals do not seek treatment, and an even smaller proportion receives FDA/EMA-approved pharmacotherapies (Grant et al., 2017; Grant et al., 2015; Mark et al., 2009; Rubinsky et al., 2015). Notably, two of the most highly regarded AD pharmacotherapies—naltrexone and acamprosate—yield suboptimal responses in approximately 40 to 70% of patients (Rosner et al., 2010a; Rosner et al., 2010b; Srisurapanont and Jarusuraisin, 2005). A recent pharmacogenomics study highlighted the association of two intergenic SNPs with specific treatment outcomes for these medications (Biernacka et al., 2021). The concept of precision medicine — tailoring treatments based on individual genetic and molecular profiles — provided the rationale for the present study ‘s hypothesis that genetic predispositions might reveal who would benefit most from OSU6162, a monoamine stabilizer showing promising preclinical and clinical results in AD settings (Feltmann et al., 2016; Fredriksson et al., 2019; Khemiri et al., 2015; Khemiri et al., 2019; Melas et al., 2021; Steensland et al., 2012).

Specifically, although OSU6162 was found to reduce alcohol craving in a 14-day randomized, double-blind, placebo-controlled trial, no treatment effects were found on alcohol intake (Khemiri et al., 2015). AD is prevalent among individuals suffering from anxiety or depression (Boschloo et al., 2011) and the results of the present study suggest that taking into consideration genetic predispositions to AD-comorbid disorders can reveal who would benefit most from OSU6162. This was supported by a robust negative correlation between anxiety PRS and drinking behaviors in AD individuals treated with OSU6162 in the original trial. Specifically, we found significant associations between anxiety factor score PRS and several clinical measures of alcohol consumption, such as the number of drinks consumed during the trial, percentage of heavy drinking days (HDD), and percentage of drinking days, including changes in blood phosphatidylethanol (PEth) levels pre- and post-treatment. A trend was also noted for changes in the percentage of drinking days pre- and post-trial. Notably, none of these relationships were observed in the placebo group.

The consistency of these associations across multiple clinical outcomes, encompassing both subjective reports and objective biomarkers, such as PEth, provides compelling evidence that underscores the potential relevance of the anxiety factor score PRS in predicting OSU6162-treatment response. Our findings also point toward the potential anxiolytic effects of OSU6162, suggesting a diminished reliance on alcohol as a coping mechanism in individuals with a genetic predisposition to anxiety (Turner et al., 2018). These anxiolytic effects may be driven by the compound ‘s monoaminergic stabilizing effects (DeGroot et al., 2020; Lawther et al., 2020) and are consistent with rodent studies demonstrating reduced alcohol intake and anxiety-like behaviors with OSU6162 treatment (Melas et al., 2021). Interestingly, however, the associations we observed were exclusively linked to the anxiety PRS based on a quantitative factor-score approach, not a binary case-control classification (Otowa et al., 2016). This emphasizes the intricate nature of anxiety phenotyping and its genetic determinants, as supported by other GWAS findings examining both continuous and binary classifications of anxiety (Levey et al., 2020). Taken together, these insights stress the importance of precise measures in psychiatric phenotyping for the advancement of precision medicine.

In the present study, the notable associations with anxiety factor score PRS were driven by a genome-wide significant threshold involving a single SNP, rs1067327. This SNP lies within a chromosomal region that implicates three primary genes, i.e., calmodulin-lysine N-methyltransferase (*CAMKMT*), prolyl endopeptidase like (*PREPL*), and solute carrier family 3 member 1 (*SLC3A1*) (Otowa et al., 2016). A PRS based on a single SNP does not capture the polygenic nature of anxiety and may not robustly predict treatment response, warranting larger and more comprehensive GWASs to develop PRSs that can be reliably utilized in clinical practice. Nonetheless, the breadth of the PRS associations with diverse clinical metrics observed in our study, particularly with an objective measure like PEth, suggests that this genetic marker may have a significant impact on the pharmacological response to OSU6162, supporting the need for biological validations of the SNP ‘s putative functional role.

Additional limitations need to be acknowledged in our study. Firstly, the analysis was conducted on a relatively small cohort of individuals (N=56), mostly of European ancestry. Consequently, the generalizability of our results to populations with diverse genetic backgrounds may be limited, highlighting the need for validation in larger and more diverse patient cohorts. Secondly, the retrospective design of our study is characteristic of early research in the field of personalized medicine for AD, where studies often retrospectively examine pharmacogenetic moderators, typically candidate SNPs, yielding results that have not been consistently replicable (Lohoff, 2020). Indeed, the retrospective approach has been shown to be a major limitation, as evidenced by pharmacogenetic biomarkers that appeared promising but later failed to demonstrate predictive power in prospective studies—such as those involving a functional SNP in the *OPRM1* gene (Anton et al., 2008; Oroszi et al., 2009; Oslin et al., 2003; Oslin et al., 2015). This underscores the imperative for prospective research to substantiate the predictive validity of genetic markers for treatment responses to interventions like OSU6162. Thirdly, our research focused solely on PRS as the predictor of treatment response, without incorporating the multiomics approaches that have been increasingly recognized as pivotal in advancing precision medicine for various diseases including asthma, cancer, infectious diseases, and metabolic disorders (Ayton et al., 2022; Hu and Jia, 2021; Logotheti et al., 2021; Ward et al., 2021). To date, only a limited number of studies have employed an omics-based strategy in AD treatment research. Notably, targeted metabolomics has been used to assess the treatment response to acamprosate, offering valuable preliminary insights (Hinton et al., 2017; Nam et al., 2015). For example, it has been shown that elevated baseline glutamate levels could predict a favorable response to acamprosate (Nam et al., 2015). Therefore, there is a compelling case for expanding precision medicine initiatives in AD to include pharmacomultiomics, aiming to uncover a broader spectrum of molecular biosignatures that could enhance the prediction of treatment outcomes.

In conclusion, although the findings of the present study are preliminary, they indicate the potential role of anxiety PRS in optimizing the use of monoaminergic stabilizers like OSU6162 for the management of alcohol use disorders. Larger clinical trials of such compounds are warranted, aiming to evaluate more comprehensive genetic data, including additional biological markers, to identify treatment responders and shed light on therapeutic mechanisms.

## Data Availability

All data produced in the present study are available upon reasonable request to the authors

## Acknowledgements

We thank Pia Steensland and late Dr. Arvid Carlsson for their pioneering work, research contributions and inspiring discussions on OSU6162 that led to the present study. We also thank the Million Veteran Program (MVP) staff, researchers, and volunteers, who have contributed to MVP (Gaziano et al., 2016), and especially participants who previously served their country in the military and now generously agreed to enroll in the study. This research is based on data from the Million Veteran Program, Office of Research and Development, Veterans Health Administration, and was supported by the Veterans Administration (VA) Million Veteran Program (MVP) award #000 (dbGAP study accession phs001672). Support by the National Bioinformatics Infrastructure Sweden (NBIS) is also gratefully acknowledged. The computations were enabled by resources provided by the National Academic Infrastructure for Supercomputing in Sweden (NAISS) at Uppsala Multidisciplinary Center for Advanced Computational Science (UPPMAX) partially funded by the Swedish Research Council through grant agreement no. 2022-06725. This work was also supported by the Royal Physiographic Society in Lund (42797, 2022; P.A.M.), the Åke Wiberg Foundation (M22-0059, 2022; P.A.M.), the Magnus Bergvall Foundation (2022-038, 2022; P.A.M.), the Sigurd and Elsa Golje Memorial Foundation (LA2022-0139; P.A.M.), and the Karolinska Institutet Research Grants (2022-01667, 2022; PAM). The funding sources were not involved in the study design or the decision to submit the results for publication.

## Author contributions

M.H.: Investigation; methodology; writing—review and editing. L.K.: Methodology; project administration; writing—review and editing. J.G.: Methodology; project administration; writing—review and editing. J.F.: Methodology; funding acquisition; project administration; resources; writing—review and editing. N.J.: Methodology; funding acquisition; project administration; resources; writing—review and editing. P.A.M.: Conceptualization; funding acquisition; methodology; project administration; resources; supervision; writing—original draft. All authors have read and agreed to the published version of the manuscript.

