## Supplementals for "Genetic liability for anxiety associates with treatment response to the monoamine stabilizer OSU6162 in alcohol dependence"

**Table S1.** Base datasets

| **Disorder or trait** | **Base study reference** | **Ancestry** | **N subjects** |
| --- | --- | --- | --- |
| Anxiety (case-control) | (Otowa et al., 2016) | European | 17,310 |
| Anxiety (factor score) |  |  | 18,186 |
| Depression | (Howard et al., 2019) | Mixed | 500,199 |
| Drinks per week | (Saunders et al., 2022) | European | 666,978 |
| Alcohol use disorder | (Kranzler et al., 2019) | European | 202,004 |
| Alcohol consumption (AUDIT-C) |  |  | 200,680 |
| Problematic alcohol use | (Zhou et al., 2020) | European | 435,563 |

AUDIT-C: Alcohol Use Disorder Identification Test-Consumption

**Table S2.** Associations between disorder or trait-PRSs and treatment (OSU6162 and placebo)

| **Disorder or trait** | **5e-08** | **0.001** | **0.05** | **0.1** | **0.2** | **0.3** | **0.4** | **0.5** | **1** |
| --- | --- | --- | --- | --- | --- | --- | --- | --- | --- |
| Alcohol consumption (AUDIT-C) | 0.704 | 0.997 | 0.704 | 0.248 | 0.248 | 0.248 | 0.248 | 0.248 | 0.248 |
| Alcohol use disorder | 0.896 | 0.896 | 0.896 | 0.899 | 0.896 | 0.896 | 0.896 | 0.896 | 0.896 |
| Anxiety (factor score) | 0.744 | 0.744 | 0.744 | 0.744 | 0.828 | 0.744 | 0.744 | 0.744 | 0.744 |
| Depression | 0.666 | 0.351 | 0.666 | 0.666 | 0.666 | 0.666 | 0.666 | 0.666 | 0.666 |
| Drinks per week | 0.432 | 0.599 | 0.432 | 0.432 | 0.432 | 0.432 | 0.432 | 0.432 | 0.432 |
| Problematic alcohol use | 0.573 | 0.361 | 0.361 | 0.361 | 0.361 | 0.361 | 0.361 | 0.361 | 0.361 |
| Anxiety (case-control) |  | 0.958 | 0.958 | 0.958 | 0.958 | 0.958 | 0.958 | 0.958 | 0.958 |

The table header lists the p-value cut-offs for each polygenic risk score (PRS) assessed.

The p-values presented in the table have been adjusted for multiple testing using the Benjamini-Hochberg method to control the false discovery rate (FDR) and are derived from t-test analyses.

AUDIT-C: Alcohol Use Disorder Identification Test-Consumption

**Table S3.** Impact of drinks-per-week PRS on clinical measures: Interaction with OSU6162 or placebo treatment

| **Clinical measures^‡^** | **5e-08** | **0.001** | **0.05** | **0.1** | **0.2** | **0.3** | **0.4** | **0.5** | **1** |
| --- | --- | --- | --- | --- | --- | --- | --- | --- | --- |
| Change % heavy drinking days | 0.966 | 0.998 | 0.861 | 0.957 | 0.985 | 0.979 | 0.984 | 0.994 | 0.935 |
| Change % drinking days | 0.966 | 0.998 | 0.861 | 0.923 | 0.985 | 0.979 | 0.984 | 0.996 | 0.935 |
| Change MADRS-S | 0.966 | 0.998 | 0.861 | 0.923 | 0.985 | 0.979 | 0.984 | 0.996 | 0.935 |
| Change PACS | 0.966 | 0.998 | 0.861 | 0.923 | 0.985 | 0.979 | 0.984 | 0.994 | 0.935 |
| Change PEth | 0.966 | 0.998 | 0.822 | 0.923 | 0.985 | 0.979 | 0.984 | 0.994 | 0.935 |
| Study drinks | 0.966 | 0.998 | 0.861 | 0.923 | 0.985 | 0.979 | 0.984 | 0.996 | 0.935 |
| Study % heavy drinking days | 0.966 | 0.998 | 0.861 | 0.923 | 0.985 | 0.979 | 0.984 | 0.994 | 0.935 |
| Study % drinking days | 0.966 | 0.998 | 0.861 | 0.923 | 0.985 | 0.979 | 0.984 | 0.994 | 0.935 |
| Craving, active cue (DAQ) | 0.966 | 0.998 | 0.708 | 0.923 | 0.985 | 0.979 | 0.984 | 0.994 | 0.935 |
| Craving, neutral cue (DAQ) | 0.966 | 0.998 | 0.861 | 0.923 | 0.985 | 0.979 | 0.984 | 0.994 | 0.935 |
| Craving, priming (DAQ) | 0.966 | 0.998 | 0.937 | 0.957 | 0.985 | 0.979 | 0.984 | 0.994 | 0.935 |
| Craving, active cue (VAS) | 0.966 | 0.998 | 0.861 | 0.923 | 0.985 | 0.979 | 0.984 | 0.994 | 0.935 |
| Craving neutral cue (VAS) | 0.966 | 0.998 | 0.861 | 0.957 | 0.985 | 0.979 | 0.984 | 0.994 | 0.935 |
| Craving priming (VAS) | 0.966 | 0.998 | 0.861 | 0.923 | 0.985 | 0.979 | 0.984 | 0.994 | 0.935 |

The table header lists the p-value cut-offs for each polygenic risk score (PRS) assessed.

The p-values presented in the table result from linear regression analyses of clinical measures based on the interaction between the anxiety factor score PRS and treatment type (OSU6162 or placebo). They have been adjusted for the false discovery rate (FDR) using the Benjamini-Hochberg method to account for multiple testing across 14 clinical measures.

^‡^Clinical measures: Change % heavy drinking days: The change in the percentage of heavy drinking days from baseline (90 days, Timeline Follow Back) to the 14-day treatment period; Change % drinking days: The change in the percentage of drinking days from baseline (90 days, Timeline Follow Back) to the 14-day treatment period; Change MADRS-S: The change in Montgomery-Åsberg Depression Self-Rating Scale (MADRS-S) scores from baseline to end of treatment (day 15); Change PACS: The change in Pennsylvania Alcohol Craving Scale (PACS) scores from baseline to end of treatment (day 15); Change PEth: The change in blood phosphatidylethanol (PEth) levels from baseline to end of treatment (day 15); Study drinks: The total number of drinks consumed during the 14-day treatment period; Study % heavy drinking days: The percentage of heavy drinking days during the 14-day treatment period; Study % drinking days: The percentage of drinking days during the 14-day treatment period; Craving (DAQ): The change in Desire for Alcohol Questionnaire (DAQ) scores immediately after the craving session compared to baseline; Craving (VAS): The change in Visual Analog Scale (VAS) scores immediately after the craving session compared to baseline.

**Table S4.** Impact of alcohol use disorder PRS on clinical measures: Interaction with OSU6162 or placebo treatment

| **Clinical measures^‡^** | **5e-08** | **0.001** | **0.05** | **0.1** | **0.2** | **0.3** | **0.4** | **0.5** | **1** |
| --- | --- | --- | --- | --- | --- | --- | --- | --- | --- |
| Change % heavy drinking days | 0.263 | 0.92 | 0.999 | 0.857 | 0.985 | 0.94 | 0.988 | 0.954 | 0.908 |
| Change % drinking days | 0.538 | 0.92 | 1 | 0.956 | 0.985 | 0.94 | 0.988 | 0.954 | 0.908 |
| Change MADRS-S | 0.252 | 0.92 | 1 | 0.857 | 0.985 | 0.94 | 0.988 | 0.954 | 0.949 |
| Change PACS | 0.538 | 0.92 | 1 | 0.874 | 0.985 | 0.94 | 0.943 | 0.954 | 0.908 |
| Change PEth | 0.24 | 0.92 | 0.999 | 0.857 | 0.985 | 0.94 | 0.988 | 0.954 | 0.949 |
| Study drinks | 0.538 | 0.92 | 0.999 | 0.857 | 0.985 | 0.94 | 0.988 | 0.954 | 0.949 |
| Study % heavy drinking days | 0.661 | 0.92 | 0.999 | 0.857 | 0.985 | 0.94 | 0.988 | 0.954 | 0.908 |
| Study % drinking days | 0.661 | 0.92 | 0.999 | 0.857 | 0.985 | 0.94 | 0.988 | 0.954 | 0.949 |
| Craving, active cue (DAQ) | 0.862 | 0.92 | 0.999 | 0.857 | 0.985 | 0.94 | 0.988 | 0.954 | 0.908 |
| Craving, neutral cue (DAQ) | 0.856 | 0.92 | 1 | 0.956 | 0.985 | 0.94 | 0.988 | 0.954 | 0.908 |
| Craving, priming (DAQ) | 0.856 | 0.92 | 0.999 | 0.857 | 0.985 | 0.94 | 0.988 | 0.954 | 0.949 |
| Craving, active cue (VAS) | 0.661 | 0.92 | 0.999 | 0.857 | 0.985 | 0.94 | 0.943 | 0.954 | 0.908 |
| Craving neutral cue (VAS) | 0.925 | 0.92 | 1 | 0.857 | 0.985 | 0.94 | 0.988 | 0.954 | 0.949 |
| Craving priming (VAS) | 0.856 | 0.92 | 1 | 0.857 | 0.985 | 0.94 | 0.988 | 0.954 | 0.908 |

The table header lists the p-value cut-offs for each polygenic risk score (PRS) assessed.

The p-values presented in the table result from linear regression analyses of clinical measures based on the interaction between the anxiety factor score PRS and treatment type (OSU6162 or placebo). They have been adjusted for the false discovery rate (FDR) using the Benjamini-Hochberg method to account for multiple testing across 14 clinical measures.

^‡^Clinical measures: Change % heavy drinking days: The change in the percentage of heavy drinking days from baseline (90 days, Timeline Follow Back) to the 14-day treatment period; Change % drinking days: The change in the percentage of drinking days from baseline (90 days, Timeline Follow Back) to the 14-day treatment period; Change MADRS-S: The change in Montgomery-Åsberg Depression Self-Rating Scale (MADRS-S) scores from baseline to end of treatment (day 15); Change PACS: The change in Pennsylvania Alcohol Craving Scale (PACS) scores from baseline to end of treatment (day 15); Change PEth: The change in blood phosphatidylethanol (PEth) levels from baseline to end of treatment (day 15); Study drinks: The total number of drinks consumed during the 14-day treatment period; Study % heavy drinking days: The percentage of heavy drinking days during the 14-day treatment period; Study % drinking days: The percentage of drinking days during the 14-day treatment period; Craving (DAQ): The change in Desire for Alcohol Questionnaire (DAQ) scores immediately after the craving session compared to baseline; Craving (VAS): The change in Visual Analog Scale (VAS) scores immediately after the craving session compared to baseline.

**Table S5.** Impact of alcohol consumption (AUDIT-C) PRS on clinical measures: Interaction with OSU6162 or placebo treatment

| **Clinical measures^‡^** | **5e-08** | **0.001** | **0.05** | **0.1** | **0.2** | **0.3** | **0.4** | **0.5** | **1** |
| --- | --- | --- | --- | --- | --- | --- | --- | --- | --- |
| Change % heavy drinking days | 0.308 | 0.841 | 0.97 | 0.681 | 0.69 | 0.717 | 0.301 | 0.356 | 0.599 |
| Change % drinking days | 0.585 | 0.995 | 0.97 | 0.752 | 0.69 | 0.717 | 0.301 | 0.292 | 0.599 |
| Change MADRS-S | 0.308 | 0.95 | 0.97 | 0.992 | 0.886 | 0.928 | 0.907 | 0.917 | 0.942 |
| Change PACS | 0.585 | 0.95 | 0.97 | 0.982 | 0.927 | 0.989 | 0.907 | 0.917 | 0.942 |
| Change PEth | 0.308 | 0.95 | 0.97 | 0.636 | 0.611 | 0.717 | 0.276 | 0.195 | 0.442 |
| Study drinks | 0.533 | 0.95 | 0.97 | 0.752 | 0.69 | 0.717 | 0.301 | 0.292 | 0.599 |
| Study % heavy drinking days | 0.875 | 0.95 | 0.97 | 0.752 | 0.69 | 0.717 | 0.301 | 0.292 | 0.599 |
| Study % drinking days | 0.741 | 0.995 | 0.97 | 0.89 | 0.69 | 0.717 | 0.301 | 0.292 | 0.599 |
| Craving, active cue (DAQ) | 0.992 | 0.95 | 0.97 | 0.982 | 0.991 | 0.928 | 0.907 | 0.917 | 0.942 |
| Craving, neutral cue (DAQ) | 0.992 | 0.442 | 0.97 | 0.982 | 0.886 | 0.928 | 0.907 | 0.917 | 0.942 |
| Craving, priming (DAQ) | 0.914 | 0.39 | 0.97 | 0.982 | 0.886 | 0.928 | 0.907 | 0.917 | 0.942 |
| Craving, active cue (VAS) | 0.533 | 0.95 | 0.97 | 0.808 | 0.886 | 0.928 | 0.907 | 0.917 | 0.942 |
| Craving neutral cue (VAS) | 0.992 | 0.995 | 0.97 | 0.982 | 0.69 | 0.814 | 0.907 | 0.917 | 0.942 |
| Craving priming (VAS) | 0.875 | 0.841 | 0.97 | 0.992 | 0.886 | 0.928 | 0.907 | 0.917 | 0.942 |

The table header lists the p-value cut-offs for each polygenic risk score (PRS) assessed.

The p-values presented in the table result from linear regression analyses of clinical measures based on the interaction between the anxiety factor score PRS and treatment type (OSU6162 or placebo). They have been adjusted for the false discovery rate (FDR) using the Benjamini-Hochberg method to account for multiple testing across 14 clinical measures.

^‡^Clinical measures: Change % heavy drinking days: The change in the percentage of heavy drinking days from baseline (90 days, Timeline Follow Back) to the 14-day treatment period; Change % drinking days: The change in the percentage of drinking days from baseline (90 days, Timeline Follow Back) to the 14-day treatment period; Change MADRS-S: The change in Montgomery-Åsberg Depression Self-Rating Scale (MADRS-S) scores from baseline to end of treatment (day 15); Change PACS: The change in Pennsylvania Alcohol Craving Scale (PACS) scores from baseline to end of treatment (day 15); Change PEth: The change in blood phosphatidylethanol (PEth) levels from baseline to end of treatment (day 15); Study drinks: The total number of drinks consumed during the 14-day treatment period; Study % heavy drinking days: The percentage of heavy drinking days during the 14-day treatment period; Study % drinking days: The percentage of drinking days during the 14-day treatment period; Craving (DAQ): The change in Desire for Alcohol Questionnaire (DAQ) scores immediately after the craving session compared to baseline; Craving (VAS): The change in Visual Analog Scale (VAS) scores immediately after the craving session compared to baseline.

**Table S6.** Impact of problematic alcohol use PRS on clinical measures: Interaction with OSU6162 or placebo treatment

| **Clinical measures^‡^** | **5e-08** | **0.001** | **0.05** | **0.1** | **0.2** | **0.3** | **0.4** | **0.5** | **1** |
| --- | --- | --- | --- | --- | --- | --- | --- | --- | --- |
| Change % heavy drinking days | 0.173 | 0.815 | 0.661 | 0.839 | 0.662 | 0.755 | 0.636 | 0.608 | 0.57 |
| Change % drinking days | 0.657 | 0.939 | 0.589 | 0.748 | 0.662 | 0.755 | 0.636 | 0.608 | 0.578 |
| Change MADRS-S | 0.173 | 0.815 | 0.589 | 0.858 | 0.697 | 0.755 | 0.788 | 0.849 | 0.943 |
| Change PACS | 0.508 | 0.815 | 0.697 | 0.748 | 0.662 | 0.755 | 0.636 | 0.608 | 0.57 |
| Change PEth | 0.0872 | 0.815 | 0.801 | 0.748 | 0.697 | 0.755 | 0.788 | 0.804 | 0.768 |
| Study drinks | 0.502 | 0.815 | 0.589 | 0.748 | 0.662 | 0.755 | 0.636 | 0.608 | 0.57 |
| Study % heavy drinking days | 0.508 | 0.815 | 0.566 | 0.748 | 0.662 | 0.755 | 0.636 | 0.608 | 0.57 |
| Study % drinking days | 0.657 | 0.815 | 0.589 | 0.748 | 0.662 | 0.755 | 0.641 | 0.608 | 0.57 |
| Craving, active cue (DAQ) | 0.761 | 0.815 | 0.589 | 0.748 | 0.763 | 0.755 | 0.636 | 0.608 | 0.57 |
| Craving, neutral cue (DAQ) | 0.838 | 0.815 | 0.292 | 0.748 | 0.662 | 0.755 | 0.636 | 0.608 | 0.57 |
| Craving, priming (DAQ) | 0.838 | 0.815 | 0.566 | 0.748 | 0.697 | 0.755 | 0.636 | 0.608 | 0.57 |
| Craving, active cue (VAS) | 0.508 | 0.815 | 0.589 | 0.855 | 0.697 | 0.755 | 0.636 | 0.608 | 0.578 |
| Craving neutral cue (VAS) | 0.761 | 0.815 | 0.589 | 0.748 | 0.838 | 0.755 | 0.67 | 0.772 | 0.794 |
| Craving priming (VAS) | 0.927 | 0.815 | 0.566 | 0.748 | 0.86 | 0.755 | 0.641 | 0.623 | 0.851 |

The table header lists the p-value cut-offs for each polygenic risk score (PRS) assessed.

The p-values presented in the table result from linear regression analyses of clinical measures based on the interaction between the anxiety factor score PRS and treatment type (OSU6162 or placebo). They have been adjusted for the false discovery rate (FDR) using the Benjamini-Hochberg method to account for multiple testing across 14 clinical measures.

^‡^Clinical measures: Change % heavy drinking days: The change in the percentage of heavy drinking days from baseline (90 days, Timeline Follow Back) to the 14-day treatment period; Change % drinking days: The change in the percentage of drinking days from baseline (90 days, Timeline Follow Back) to the 14-day treatment period; Change MADRS-S: The change in Montgomery-Åsberg Depression Self-Rating Scale (MADRS-S) scores from baseline to end of treatment (day 15); Change PACS: The change in Pennsylvania Alcohol Craving Scale (PACS) scores from baseline to end of treatment (day 15); Change PEth: The change in blood phosphatidylethanol (PEth) levels from baseline to end of treatment (day 15); Study drinks: The total number of drinks consumed during the 14-day treatment period; Study % heavy drinking days: The percentage of heavy drinking days during the 14-day treatment period; Study % drinking days: The percentage of drinking days during the 14-day treatment period; Craving (DAQ): The change in Desire for Alcohol Questionnaire (DAQ) scores immediately after the craving session compared to baseline; Craving (VAS): The change in Visual Analog Scale (VAS) scores immediately after the craving session compared to baseline.

**Table S7.** Impact of anxiety (case-control) PRS on clinical measures: Interaction with OSU6162 or placebo treatment

| **Clinical measures^‡^** | **0.001** | **0.05** | **0.1** | **0.2** | **0.3** | **0.4** | **0.5** | **1** |
| --- | --- | --- | --- | --- | --- | --- | --- | --- |
| Change % heavy drinking days | 0.908 | 0.278 | 0.68 | 0.471 | 0.417 | 0.643 | 0.523 | 0.532 |
| Change % drinking days | 0.908 | 0.234 | 0.551 | 0.35 | 0.417 | 0.642 | 0.467 | 0.532 |
| Change MADRS-S | 0.945 | 0.19 | 0.221 | 0.35 | 0.417 | 0.643 | 0.523 | 0.589 |
| Change PACS | 0.908 | 0.278 | 0.284 | 0.35 | 0.31 | 0.642 | 0.467 | 0.532 |
| Change PEth | 0.908 | 0.607 | 0.699 | 0.612 | 0.625 | 0.802 | 0.523 | 0.532 |
| Study drinks | 0.908 | 0.278 | 0.605 | 0.612 | 0.835 | 0.959 | 0.792 | 0.875 |
| Study % heavy drinking days | 0.908 | 0.278 | 0.699 | 0.777 | 0.835 | 0.959 | 0.792 | 0.875 |
| Study % drinking days | 0.908 | 0.278 | 0.551 | 0.605 | 0.625 | 0.802 | 0.523 | 0.589 |
| Craving, active cue (DAQ) | 0.908 | 0.19 | 0.221 | 0.35 | 0.417 | 0.642 | 0.523 | 0.532 |
| Craving, neutral cue (DAQ) | 0.945 | 0.671 | 0.853 | 0.982 | 0.835 | 0.921 | 0.792 | 0.971 |
| Craving, priming (DAQ) | 0.908 | 0.855 | 0.853 | 0.982 | 0.835 | 0.921 | 0.672 | 0.589 |
| Craving, active cue (VAS) | 0.908 | 0.396 | 0.699 | 0.605 | 0.417 | 0.642 | 0.523 | 0.532 |
| Craving neutral cue (VAS) | 0.908 | 0.8 | 0.699 | 0.605 | 0.417 | 0.642 | 0.523 | 0.532 |
| Craving priming (VAS) | 0.945 | 0.8 | 0.914 | 0.982 | 0.835 | 0.921 | 0.668 | 0.589 |

The table header lists the p-value cut-offs for each polygenic risk score (PRS) assessed.

The p-values presented in the table result from linear regression analyses of clinical measures based on the interaction between the anxiety factor score PRS and treatment type (OSU6162 or placebo). They have been adjusted for the false discovery rate (FDR) using the Benjamini-Hochberg method to account for multiple testing across 14 clinical measures.

^‡^Clinical measures: Change % heavy drinking days: The change in the percentage of heavy drinking days from baseline (90 days, Timeline Follow Back) to the 14-day treatment period; Change % drinking days: The change in the percentage of drinking days from baseline (90 days, Timeline Follow Back) to the 14-day treatment period; Change MADRS-S: The change in Montgomery-Åsberg Depression Self-Rating Scale (MADRS-S) scores from baseline to end of treatment (day 15); Change PACS: The change in Pennsylvania Alcohol Craving Scale (PACS) scores from baseline to end of treatment (day 15); Change PEth: The change in blood phosphatidylethanol (PEth) levels from baseline to end of treatment (day 15); Study drinks: The total number of drinks consumed during the 14-day treatment period; Study % heavy drinking days: The percentage of heavy drinking days during the 14-day treatment period; Study % drinking days: The percentage of drinking days during the 14-day treatment period; Craving (DAQ): The change in Desire for Alcohol Questionnaire (DAQ) scores immediately after the craving session compared to baseline; Craving (VAS): The change in Visual Analog Scale (VAS) scores immediately after the craving session compared to baseline.

**Table S8.** Impact of depression PRS on clinical measures: Interaction with OSU6162 or placebo treatment

| **Clinical measures^‡^** | **5e-08** | **0.001** | **0.05** | **0.1** | **0.2** | **0.3** | **0.4** | **0.5** | **1** |
| --- | --- | --- | --- | --- | --- | --- | --- | --- | --- |
| Change % heavy drinking days | 0.977 | 0.594 | 0.848 | 0.71 | 0.772 | 0.657 | 0.622 | 0.686 | 0.669 |
| Change % drinking days | 0.977 | 0.515 | 0.678 | 0.675 | 0.772 | 0.657 | 0.622 | 0.686 | 0.669 |
| Change MADRS-S | 0.977 | 0.069 | 0.605 | 0.71 | 0.859 | 0.749 | 0.705 | 0.722 | 0.707 |
| Change PACS | 0.935 | 0.456 | 0.848 | 0.822 | 0.772 | 0.648 | 0.622 | 0.542 | 0.428 |
| Change PEth | 0.935 | 0.515 | 0.729 | 0.854 | 0.859 | 0.657 | 0.622 | 0.686 | 0.669 |
| Study drinks | 0.977 | 0.515 | 0.371 | 0.675 | 0.445 | 0.289 | 0.247 | 0.297 | 0.249 |
| Study % heavy drinking days | 0.977 | 0.515 | 0.371 | 0.675 | 0.452 | 0.289 | 0.247 | 0.297 | 0.249 |
| Study % drinking days | 0.977 | 0.515 | 0.371 | 0.566 | 0.445 | 0.289 | 0.247 | 0.297 | 0.249 |
| Craving, active cue (DAQ) | 0.67 | 0.515 | 0.371 | 0.258 | 0.0502 | 0.0604 | 0.125 | 0.0743 | **0.0401*** |
| Craving, neutral cue (DAQ) | 0.935 | 0.515 | 0.873 | 0.923 | 0.859 | 0.657 | 0.622 | 0.686 | 0.669 |
| Craving, priming (DAQ) | 0.977 | 0.515 | 0.371 | 0.464 | 0.255 | 0.247 | 0.125 | 0.114 | 0.0724 |
| Craving, active cue (VAS) | 0.67 | 0.515 | 0.678 | 0.71 | 0.772 | 0.657 | 0.705 | 0.718 | 0.751 |
| Craving neutral cue (VAS) | 0.935 | 0.515 | 0.848 | 0.854 | 0.851 | 0.657 | 0.705 | 0.686 | 0.669 |
| Craving priming (VAS) | 0.67 | 0.75 | 0.873 | 0.854 | 0.859 | 0.839 | 0.735 | 0.722 | 0.707 |

The table header lists the p-value cut-offs for each polygenic risk score (PRS) assessed.

The p-values presented in the table result from linear regression analyses of clinical measures based on the interaction between the anxiety factor score PRS and treatment type (OSU6162 or placebo). They have been adjusted for the false discovery rate (FDR) using the Benjamini-Hochberg method to account for multiple testing across 14 clinical measures, with significance denoted as *p<0.05.

^‡^Clinical measures: Change % heavy drinking days: The change in the percentage of heavy drinking days from baseline (90 days, Timeline Follow Back) to the 14-day treatment period; Change % drinking days: The change in the percentage of drinking days from baseline (90 days, Timeline Follow Back) to the 14-day treatment period; Change MADRS-S: The change in Montgomery-Åsberg Depression Self-Rating Scale (MADRS-S) scores from baseline to end of treatment (day 15); Change PACS: The change in Pennsylvania Alcohol Craving Scale (PACS) scores from baseline to end of treatment (day 15); Change PEth: The change in blood phosphatidylethanol (PEth) levels from baseline to end of treatment (day 15); Study drinks: The total number of drinks consumed during the 14-day treatment period; Study % heavy drinking days: The percentage of heavy drinking days during the 14-day treatment period; Study % drinking days: The percentage of drinking days during the 14-day treatment period; Craving (DAQ): The change in Desire for Alcohol Questionnaire (DAQ) scores immediately after the craving session compared to baseline; Craving (VAS): The change in Visual Analog Scale (VAS) scores immediately after the craving session compared to baseline.

**Figure S1**

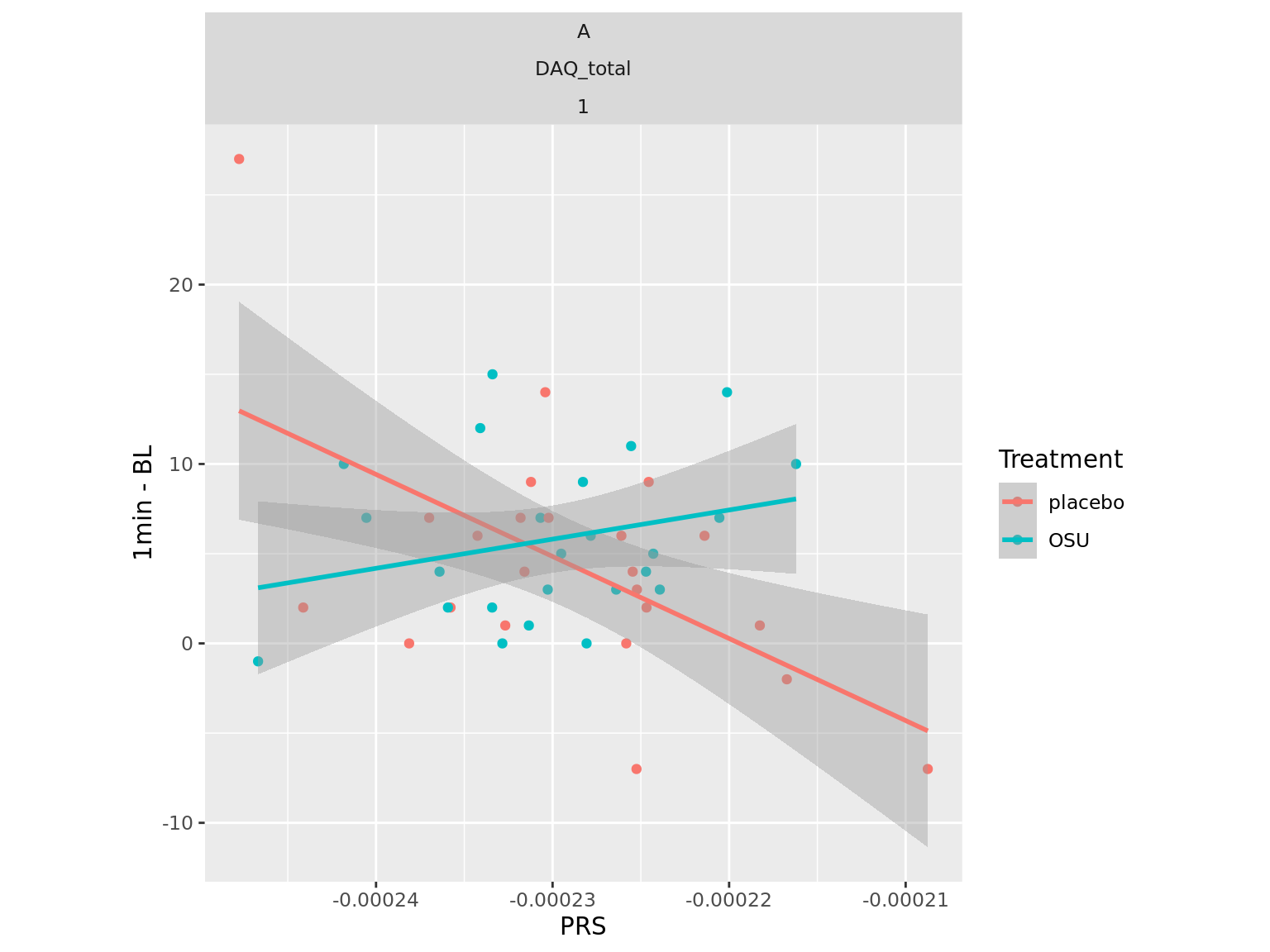

**Fig S1.** **Correlation between depression PRS and craving in alcohol dependent individuals on OSU6162 or placebo.** Depression PRS correlated negatively with craving in the placebo group. No such effect was present in the OSU6162 group. A: active (alcohol specific) cue session; DAQ_total (1min – BL): The change in Desire for Alcohol Questionnaire (DAQ) scores immediately after the craving session compared to baseline; 1: The p-value cutoff for PRS. Treatment groups are indicated by color: red for placebo and teal for OSU6162 (OSU).

Saunders, G.R.B., Wang, X., Chen, F., Jang, S.K., Liu, M., Wang, C., Gao, S., Jiang, Y., Khunsriraksakul, C., Otto, J.M., Addison, C., Akiyama, M., Albert, C.M., Aliev, F., Alonso, A., Arnett, D.K., Ashley-Koch, A.E., Ashrani, A.A., Barnes, K.C., Barr, R.G., Bartz, T.M., Becker, D.M., Bielak, L.F., Benjamin, E.J., Bis, J.C., Bjornsdottir, G., Blangero, J., Bleecker, E.R., Boardman, J.D., Boerwinkle, E., Boomsma, D.I., Boorgula, M.P., Bowden, D.W., Brody, J.A., Cade, B.E., Chasman, D.I., Chavan, S., Chen, Y.I., Chen, Z., Cheng, I., Cho, M.H., Choquet, H., Cole, J.W., Cornelis, M.C., Cucca, F., Curran, J.E., de Andrade, M., Dick, D.M., Docherty, A.R., Duggirala, R., Eaton, C.B., Ehringer, M.A., Esko, T., Faul, J.D., Fernandes Silva, L., Fiorillo, E., Fornage, M., Freedman, B.I., Gabrielsen, M.E., Garrett, M.E., Gharib, S.A., Gieger, C., Gillespie, N., Glahn, D.C., Gordon, S.D., Gu, C.C., Gu, D., Gudbjartsson, D.F., Guo, X., Haessler, J., Hall, M.E., Haller, T., Harris, K.M., He, J., Herd, P., Hewitt, J.K., Hickie, I., Hidalgo, B., Hokanson, J.E., Hopfer, C., Hottenga, J., Hou, L., Huang, H., Hung, Y.J., Hunter, D.J., Hveem, K., Hwang, S.J., Hwu, C.M., Iacono, W., Irvin, M.R., Jee, Y.H., Johnson, E.O., Joo, Y.Y., Jorgenson, E., Justice, A.E., Kamatani, Y., Kaplan, R.C., Kaprio, J., Kardia, S.L.R., Keller, M.C., Kelly, T.N., Kooperberg, C., Korhonen, T., Kraft, P., Krauter, K., Kuusisto, J., Laakso, M., Lasky-Su, J., Lee, W.J., Lee, J.J., Levy, D., Li, L., Li, K., Li, Y., Lin, K., Lind, P.A., Liu, C., Lloyd-Jones, D.M., Lutz, S.M., Ma, J., Magi, R., Manichaikul, A., Martin, N.G., Mathur, R., Matoba, N., McArdle, P.F., McGue, M., McQueen, M.B., Medland, S.E., Metspalu, A., Meyers, D.A., Millwood, I.Y., Mitchell, B.D., Mohlke, K.L., Moll, M., Montasser, M.E., Morrison, A.C., Mulas, A., Nielsen, J.B., North, K.E., Oelsner, E.C., Okada, Y., Orru, V., Palmer, N.D., Palviainen, T., Pandit, A., Park, S.L., Peters, U., Peters, A., Peyser, P.A., Polderman, T.J.C., Rafaels, N., Redline, S., Reed, R.M., Reiner, A.P., Rice, J.P., Rich, S.S., Richmond, N.E., Roan, C., Rotter, J.I., Rueschman, M.N., Runarsdottir, V., Saccone, N.L., Schwartz, D.A., Shadyab, A.H., Shi, J., Shringarpure, S.S., Sicinski, K., Skogholt, A.H., Smith, J.A., Smith, N.L., Sotoodehnia, N., Stallings, M.C., Stefansson, H., Stefansson, K., Stitzel, J.A., Sun, X., Syed, M., Tal-Singer, R., Taylor, A.E., Taylor, K.D., Telen, M.J., Thai, K.K., Tiwari, H., Turman, C., Tyrfingsson, T., Wall, T.L., Walters, R.G., Weir, D.R., Weiss, S.T., White, W.B., Whitfield, J.B., Wiggins, K.L., Willemsen, G., Willer, C.J., Winsvold, B.S., Xu, H., Yanek, L.R., Yin, J., Young, K.L., Young, K.A., Yu, B., Zhao, W., Zhou, W., Zollner, S., Zuccolo, L., andMe Research, T., Biobank Japan, P., Batini, C., Bergen, A.W., Bierut, L.J., David, S.P., Gagliano Taliun, S.A., Hancock, D.B., Jiang, B., Munafo, M.R., Thorgeirsson, T.E., Liu, D.J., Vrieze, S., 2022. Genetic diversity fuels gene discovery for tobacco and alcohol use. Nature 612(7941), 720-724.

Zhou, H., Sealock, J.M., Sanchez-Roige, S., Clarke, T.K., Levey, D.F., Cheng, Z., Li, B., Polimanti, R., Kember, R.L., Smith, R.V., Thygesen, J.H., Morgan, M.Y., Atkinson, S.R., Thursz, M.R., Nyegaard, M., Mattheisen, M., Borglum, A.D., Johnson, E.C., Justice, A.C., Palmer, A.A., McQuillin, A., Davis, L.K., Edenberg, H.J., Agrawal, A., Kranzler, H.R., Gelernter, J., 2020. Genome-wide meta-analysis of problematic alcohol use in 435,563 individuals yields insights into biology and relationships with other traits. Nat Neurosci 23(7), 809-818.
